# Comparative Validation of BSE and EACVI/ASE Guidelines for Estimating Filling Pressure: Proposal of an Algorithm

**DOI:** 10.1101/2025.07.09.25331065

**Authors:** Amer Barakat, Omar Alkassem, Ahmad Rasheed Alsaadi

## Abstract

**Background:** Accurate noninvasive assessment of left ventricular (LV) filling pressure is pivotal in evaluating diastolic function. While both the EACVI/ASE 2016 and BSE 2024 guidelines provide structured echocardiographic criteria, their diagnostic agreement and alignment with invasive measurements remain unclear.

**Objectives:** To compare the diagnostic performance of BSE 2024 versus EACVI/ASE 2016 guidelines against invasively measured LV end-diastolic pressure (LVEDP) and LV pre-A pressure, and to propose and validate a complementary diagnostic algorithm to EACVI/ASE 2016 incorporating left atrial strain and pulmonary venous flow.

**Methods:** This prospective, multicenter study included 138 patients referred for elective left heart catheterization. Echocardiographic diastolic function was classified using both guideline systems. A novel complementary algorithm integrating LA reservoir strain (LARs), pulmonary venous S/D ratio, and Ar-A duration was evaluated. All classifications were validated against invasive hemodynamic data.

**Results:** BSE 2024 reduced indeterminate classifications (0.7%) but overclassified diastolic dysfunction (90.6%) compared with EACVI/ASE 2016 (39.9%). Both guidelines showed moderate agreement with invasive standards (AUC < 0.70). The proposed algorithm demonstrated superior diagnostic accuracy (AUC up to 0.82). No statistically difference between both guidelines for the assessment of EF preserved/reduced.

**Conclusions:** While BSE 2024 improves classification clarity, both guidelines show limited diagnostic accuracy compared to invasive measurements. A complementary algorithm based on EACVI/ASE 2016—enhanced by LARs and pulmonary venous flow—achieves significantly better performance and may guide future diagnostic strategies.

## 1. Introduction

Left ventricular (LV) diastolic dysfunction [1] is a central pathophysiological mechanism in patients with heart failure (HF), representing a diagnostic and therapeutic challenge in clinical cardiology. Accurate noninvasive assessment of LV filling pressures is therefore essential, and echocardiography, particularly Doppler-based techniques, remains the first-line modality recommended by major societies.

In 2016, the EACVI/ASE guidelines [1] proposed a multi-parametric approach relying on parameters such as E/e′ ratio, septal and lateral e′ velocities, left atrial volume index (LAVI), and tricuspid regurgitation velocity (TR). However, several validation studies have shown only moderate concordance between guideline-based classification and invasive hemodynamic reference standards (e.g., LV end-diastolic pressure LVEDP [2]).

Recently, the British Society of Echocardiography (BSE) released its 2024 update [3], which simplifies some classification criteria and emphasizes practical thresholds. Nevertheless, real-world validation of these new criteria against invasive data is lacking.

Over the past decade, left atrial (LA) strain, derived from speckle-tracking echocardiography, has emerged as a more sensitive marker of elevated LV filling pressure, often outperforming conventional diastolic indices [5, 6]. Atrial dysfunction, reflected by reduced reservoir strain (<18%), correlates with both elevated LVEDP and adverse outcomes in HF. Consequently, several recent reviews and expert consensus documents have advocated for its incorporation into diagnostic algorithms [4, 5].

Despite these advancements, significant knowledge gaps remain:

– There is no existing comparison between BSE 2024 [3] and EACVI/ASE 2016 [1] using invasive LVEDP and LV pre-A as gold standards.
– The diagnostic agreement between these guidelines is poorly understood in clinical practice.
– Importantly, no data exist for such comparisons in Middle Eastern or Syrian populations.

This multicenter prospective study addresses these gaps by:

1. Comparing BSE 2024 and EACVI/ASE 2016 diastolic function classifications against invasive LVEDP and LV pre-A pressure.
2. Proposing and validating a complementary algorithm based on EACVI/ASE 2016 integrating LA strain LARs, pulmonary venous S/D and Ar-A duration, aiming to improve diagnostic accuracy in patients who need LV diastolic function and filling pressure assessment.

## 2. Methods

### 2.1 Study Design and Population

This was a prospective, multicenter, observational study conducted across three tertiary cardiology centers in Syria (Damascus University Hospitals) between January 2023 and January 2024. A total of 138 adult patients (age ≥18 years) referred for left heart catheterization were enrolled. Patients were classified according to preserved or reduced ejection fraction based on echocardiographic measurements.

Patients with moderate/severe valvular heart disease, atrial fibrillation, prior mitral valve surgery, or poor acoustic windows were excluded. The study was approved by local institutional review boards, and informed consent was obtained from all patients.

### 2.2 Echocardiographic Evaluation

All echocardiographic studies were performed immediately prior to left heart catheterization, using commercial ultrasound machines (GE Vivid, Philips EPIQ). Certified echocardiographers blinded to invasive measurements acquired and analyzed the data.

Diastolic function and LV filling pressures were assessed using:

– EACVI/ASE 2016 [1] guidelines
– BSE 2024 [3] recommendations

The echocardiographic parameters included:

– 2D and M mode EF%
– Mitral inflow velocities (E, A, E/A)
– Tissue Doppler imaging (septal and lateral e′, averaged for E/e′)
– Left atrial volume index (LAVI)
– Peak TR velocity
– Pulmonary venous flow (S, D, A reversal)
– Ar-A duration difference
– Left atrial reservoir strain (LARs) using 2D speckle-tracking

### 2.3 Invasive Hemodynamic Assessment

Left heart catheterization was performed using femoral or radial access with fluid-filled pigtail catheters. Measurements included:

– LV end-diastolic pressure (LVEDP)
– LV pre-A pressure (immediately before atrial contraction)

Hemodynamic thresholds were defined as:

– LVEDP >15 mmHg
– LV pre-A ≥12 mmHg and another cut-off ≥15 mmHg for further assessment. Operators were blinded to echo results.

### 2.4 Proposed Algorithm

This study developed a supplemental algorithm based on the EACVI/ASE 2016 [1] guidelines. For patients categorized as ‘normal’ or ‘indeterminate’ filling pressure by EACVI/ASE 2016 but with ongoing clinical suspicion, the algorithm was applied as follow ( Figure: proposed algorithm)s:

1. **Left Atrial Reservoir Strain (LARs):**

1. ≥24% → Normal filling pressure (per BSE 2024 [3])
2. <18% → Elevated filling pressure
3. 18–24% → Proceed to step 2
2. **Pulmonary venous S/D ratio and Ar-A duration difference:**

1. If at least one positive of S/D <1 or Ar-A ≥30 ms→ Elevated filling pressure.
2. If both are normal → Normal filling pressure
3. Other possibilities (at least one missing) → Indeterminate filling pressure.

### 2.5 Statistical Analysis

Continuous data are presented as mean ± SD and compared using t-tests or ANOVA. Categorical data were compared using chi-square or Fisher’s exact test. Diagnostic performance was evaluated using ROC curves, AUC analysis with DeLong test, and diagnostic indices (sensitivity, specificity, PPV, NPV, F1 score). Agreement was assessed with Cohen’s kappa. A p-value <0.05 was considered significant. Analyses were performed using SPSS v25 and MedCalc v20.

## 3. Results

### 3.1 Study Population

The study included 138 patients (mean age 55.2 ± 8.7 years, 44.9% female). 72.5% of patients had preserved EF, with a prevalence of hypertension (54.3%), diabetes (53.6%), and ischemic heart disease (48.6%). Average LAVI was 22.5 ± 9.2 mL/m², and the mean LVEDP was 19.4 ± 7.5 mmHg; mean LV pre-A was 12.7 ± 5.7 mmHg. Full baseline characteristics are summarized in Table 1.

**Table 1:** Study population description Clinical History, Medications, and Symptoms.

| Data | Mean ±SD | Data | (%) |
| --- | --- | --- | --- |
| Age (years) | 55.17 ± 8.65 | Female | 44.9 % |
| BSA (m<sup>2</sup>) | 1.87 ± 0.16 | HTN | 54.3 % |
| LAVI (ml/m<sup>2</sup>) | 22.52 ± 9.24 | DM | 53.6 % |
| EF (%) | 55 ± 9 | IHD | 48.6 % |
| Lateral e' (cm/s) | 10.83 ± 3.12 | EF preserved patients | 72.5 % |
| Septal e' (cm/s) | 7.78 ± 2.74 | Chronic Kidney Disease (eGFR <60) | (20.3%) |
| AV E/e' | 8.54 ± 4.16 | Beta-blockers | (64.5%) |
| E/A | 1.66 ± 6.48 | ACEi/ARB | (58.7%) |
| TR (cm/s) | 1.72 ± 0.86 | Diuretics | (43.5%) |
| S/D | 1.0 ± 0.38 | Statins | (52.9%) |
| Ar-A (ms) | 20.77 ± 28.33 | Dyspnea (NYHA ≥ II) | (66.7%) |
| LARs (%) | 27.90 ± 10.61 | Orthopnea | (22.5%) |
| LVEDP mmHg | 19.37 ± 7.52 | Fatigue | (34.8%) |
| LV pre-A mmHg | 12.66 ± 5.74 | | |

BMI: body mass index, BSA: body surface area, LAVI: left atrial volume indexed, EF: ejection fraction, e': tissue velocity, E/A Mitral early to late flow ratio, E/e': averaged early mitral inflow to averaged tissue velocity ratio, TR: Tricuspid regurgitation, EDT: deceleration time, S/D: pulmonary s to d waves ratio, LVEDP: left ventricular end diastolic pressure and LV pre-A. HTN = Hypertension, DM = Diabetes Mellitus, IHD = Ischemic Heart Disease, EF: Ejection fraction, ACEi = Angiotensin-Converting Enzyme inhibitor; ARB = Angiotensin II Receptor Blocker; NYHA = New York Heart Association functional class.

### 3.2 Hemodynamic Correlation and Method Agreement

#### 3.2.1 Echo vs Catheter Measurements

There were no statistically significant differences in key hemodynamic parameters (HR, SBP, DBP, MAP) between echo and catheter data (p > 0.4 for all, Table 2).

**Table 2:**
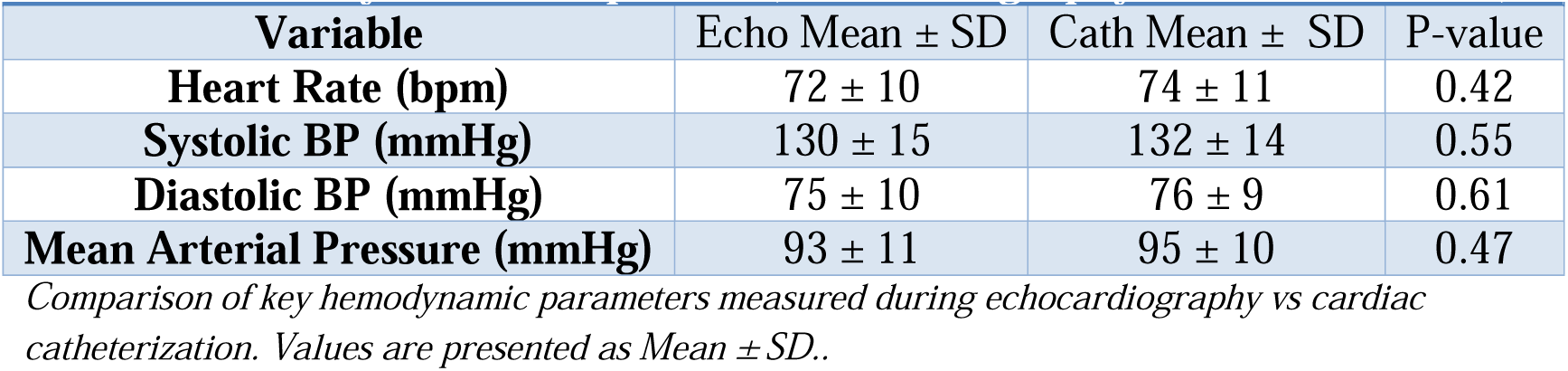
Hemodynamic Comparison (Echocardiography vs Catheterization)

#### 3.2.2 Guideline-Based Diastolic Classification

Compared to EACVI/ASE 2016 [1], the BSE 2024 [3] guideline yielded significantly fewer indeterminate cases (0.7% vs 2.9%, p < 0.001) and a higher rate of impaired classification (90.6% vs 39.9%) (Table 3).

**Table 3.** Comparison of Diastolic Function Classification Between BSE 2024 and EACVI 2016.

2 Comparison of key hemodynamic parameters measured during echocardiography vs cardiac 3 catheterization. Values are presented as Mean ± SD..
|  |  |  |  |  |
| --- | --- | --- | --- | --- |
| Guideline | (Indeterminate) | (Normal) | (Impaired) |  |
| BSE 2024 | 1 (0.7%) | 12 (8.7%) | 125 (90.6%) |  |
| EACVI/ASE 2016 | 4 (2.9%) | 79 (57.2%) | 55 (39.9%) | Grade I: 21.7%<br>Grade II: 12.3%<br>Grade III: 5.8% |
| P-value | < 0.001 | < 0.001 | < 0.001 |  |

P-value calculated using Chi-square test. BSE = British Society of Echocardiography 2024; EACVI = European Association of Cardiovascular Imaging and ASE: American Society of Cardiology

Cross-tabulation revealed moderate agreement between the two classifications (Cohen’s kappa = 0.65), with discordance in 21.7% of cases (Table 9 supplementary).

### 3.3 Invasive Pressure Comparison Across Classifications

#### 3.3.1 Stratification by Function Groups

Patients classified as having impaired diastolic function had significantly higher mean LVEDP and LV pre-A under both guidelines (p < 0.05), with larger pressure gradients under EACVI (Table 4).

**Table 4:** Comparison of Invasive Pressures by Diastolic Function Classification (EACVI vs BSE)

4 P-value calculated using Chi-square test. BSE = British Society of Echocardiography 2024; EACVI = 5 European Association of Cardiovascular Imaging and ASE: American Society of Cardiology
|  |  |  |  |  |
| --- | --- | --- | --- | --- |
| Guidelines | Group | Normal | Impaired diastolic function | P-value (comparison) |
| EACVI/ASE 2016 | Mean LVEDP ±SD (mmHg) | 16.2 ± 5.6 | 23.3 ± 8.1 | ≤0.00001 |
|  | Mean LV pre-A ± SD (mmHg) | 10.5 ± 4.3 | 15.7 ± 5.1 | ≤0.00001 |
| BSE 2024 | Mean LVEDP ±SD (mmHg) | 14.0 ± 5.6 | 19.8 ± 7.6 | p=0.01583 |
|  | Mean LV pre-A ± SD (mmHg) | 9.5 ± 3.2 | 13.0 ± 5.4 | p=0.03798 |

Cross-method Comparison Within Same Function Group
| Same Function Group | LVEDP: EACVI vs BSE | LV pre-A: EACVI vs BSE |
| --- | --- | --- |
| Normal | p=0.21684 | p=0.46177 |
| Impaired diastolic function | p=0.00787 | p=0.00299 |

*BSE = British Society of Echocardiography; EACVI = European Association of Cardiovascular Imaging,* *ASE: American Society of Cardiology. LVEDP: left ventricular end diastolic pressure..*

Further stratification by EACVI grades showed stepwise increases in LVEDP and LV pre-A with worsening grades but not between grade 2 and grade three (Table 10 supplementary).

### 3.4 Diagnostic Performance and ROC Analysis

#### 3.4.1 ROC Performance

When predicting elevated LV filling pressures:

– EACVI/ASE 2016 [1] slightly outperformed BSE 2024 [3] for all thresholds but with modest AUCs (max AUC = 0.68) (Table 5).
– BSE had higher specificity but markedly lower sensitivity.

**Table 5:** ROC Analysis of BSE and EACVI in Predicting LV pre-A > 12 mmHg, LV pre-A > 15 mmHg and LVEDP>15 mmHg.

|  |  |  |  |  |  |  |  |
| --- | --- | --- | --- | --- | --- | --- | --- |
| Pressure wave cut-off | Recommendation | AUC | Sensitivity | Specificity | PPV | NPV | P-value AUCs |
| LV pre-A > 12 mmHg | BSE 2024 | 0.645 | 0.316 | 0.974 | 0.900 | 0.655 | 0.511 |
|  | EACVI/ASE 2016 | 0.680 | 0.386 | 0.974 | 0.917 | 0.679 |  |
| LV pre-A>15 mmHg | BSE 2024 | 0.603 | 0.293 | 0.913 | 0.600 | 0.743 | 0.511 |
|  | EACVI/ASE 2016 | 0.652 | 0.390 | 0.913 | 0.667 | 0.771 |  |
| LVEDP> 15 mmHg | BSE 2024 | 0.587 | 0.221 | 0.953 | 0.895 | 0.406 | 0.499 |
|  | EACVI/ASE 2016 | 0.613 | 0.273 | 0.953 | 0.913 | 0.423 |  |

*BSE = British Society of Echocardiography, EACVI = European Association of Cardiovascular Imaging,* *ASE: American Society of Cardiology. LVEDP: left ventricular end diastolic pressure. AUC: Area under* *the curve, NPV: negative predictive value, PPV: positive predictive value. Both recommendations* *demonstrated significant associations with invasive pressure (p < 0.001).*

#### 3.4.2 Proposed Algorithm Superiority

The proposed algorithm integrating LARs, pulmonary S/D and Ar-A duration demonstrated:

– AUC = 0.82 (LV pre-A ≥12 mmHg)
– AUC = 0.76 (LV pre-A ≥15 mmHg)
– AUC = 0.80 (LVEDP [2] >15 mmHg)

This significantly outperformed both EACVI and BSE (p < 0.001 for all, Tables 7,8).

### 3.5 Subgroup Analysis by EF For preserved EF

– Improvements were smaller but consistent (e.g., AUC = 0.77 vs 0.68) all P value of AUCs comparison were > 0.05 (Table 13 supplementary).

For patients with reduced EF:

– The proposed algorithm showed the highest AUCs (up to 0.89), vs <0.65 for both guidelines, all P value of AUCs comparison were > 0.05 (Table 8).

### 3.6 Agreement in Filling Pressure Classification

The proposed algorithm classified 34.8% as elevated pressure, compared to 15.9% (BSE) and 18.1% (EACVI). Agreement between BSE and EACVI was strong (Cohen’s kappa = 0.837, Table 12 supplementary), but both diverged from the proposed method. Overview of distributions is shown in Table 6 and comparison cross-tabulations in Table 11 supplementary. The proposed method had Indeterminate portion of (15.2%).

**Table 6:**
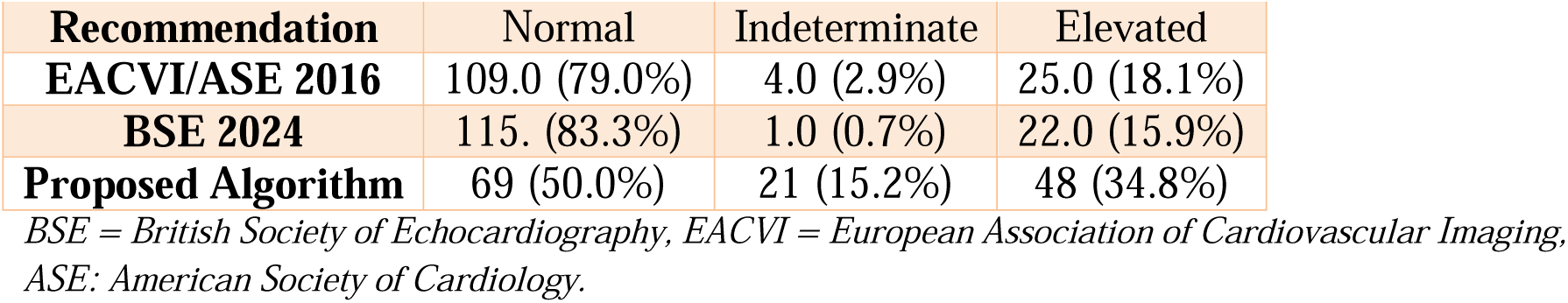
Distribution of Pressure Categories by Recommendation.

## 4. Discussion

This study provides a comprehensive validation of the recently published BSE 2024 [3] guideline compared to the widely used EACVI/ASE 2016 [1] recommendations, using invasive pressure measurements as reference standards. The findings demonstrate several key observations with important clinical implications.

First, the BSE 2024 [3] guideline significantly reduces the proportion of indeterminate diastolic function classifications compared to EACVI/ASE 2016 [1] (0.7% vs 2.9%). This supports the updated guideline’s practicality and its potential to enhance clinical decision-making by providing clearer categorization.

Second, the EACVI/ASE 2016 [1] algorithm yielded slightly better diagnostic performance in ROC analyses across all pressure cutoffs, albeit with relatively modest AUC values (<0.70) with no statistically difference between them. These findings suggest that BSE 2024 may come at the cost of sensitivity, particularly for identifying early or borderline elevations in LV filling pressure but in general statistically non inferior of EACVI/ASE 2016.

Third, the proposed diagnostic algorithm, which incorporates LA reservoir strain, pulmonary venous S/D ratio, and Ar-A duration difference, demonstrated superior performance (AUCs up to 0.82). Importantly, this algorithm maintained high diagnostic value across both preserved and reduced EF subgroups. This reinforces the growing recognition of LA strain as a sensitive and reliable indicator of elevated LV filling pressures, and its integration into routine diastolic evaluation appears well justified.

While EACVI/ASE 2016 and BSE 2024 have same 4 major echocardiographic parameters (Av E/è, TR, LAVI and è) adding the same additional criteria in BSE 2024 (S/D, Ar-A and LARs) as a complementary to EACVI/ASE 2016 showed higher diagnostic performance than both current guidelines, this highlights the importance of the combination of same criteria among different algorithms.

Looking more closely at the classification agreement (Table 11 supplementary), the strong concordance between BSE 2024 [3] and EACVI 2016 is numerically represented by 108 cases concordantly classified as normal and 19 as elevated, with only 6 discordant cases. Despite this agreement, the proposed algorithm shifted classification in 21–25% of patients, reclassifying several borderline EACVI-normal cases into the elevated pressure category based on LARs and venous metrics (Table 6).

Quantitatively, the proposed algorithm achieved superior predictive power in terms of NPV (0.698 vs 0.423 for EACVI) and comparable PPV (0.875 vs 0.913) (Table 7).

**Table 7:** ROC Analysis of proposed algorithm in Predicting LV pre-A > 12 mmHg, LV pre-A > 15 mmHg and LVEDP>15 mmHg.

|  |  |  |  |  |  |  |  |  |
| --- | --- | --- | --- | --- | --- | --- | --- | --- |
| Proposed algorithm | Pressure wave cut-off | AUC | Sensitivity | Specificity | NPV | PPV | P-value AUCs Proposed vs EACVI/ASE 2016 | P-value AUCs Proposed vs BSE 2024 |
|  | LV pre-A > 12 mmHg | 0.821 | 0.833 | 0.809 | 0.859 | 0.776 | 0.001 | <0.0001 |
|  | LV pre-A>15 mmHg | 0.760 | 0.825 | 0.695 | 0.891 | 0.569 | 0.018 | <0.0001 |
|  | LVEDP>15 mmHg | 0.797 | 0.754 | 0.841 | 0.698 | 0.875 | <0.0001 | <0.0001 |

BSE = British Society of Echocardiography, EACVI = European Association of Cardiovascular Imaging, ASE: American Society of Cardiology. LVEDP: left ventricular end diastolic pressure. AUC: Area under the curve, NPV: negative predictive value, PPV: positive predictive value.

**Table 8:** ROC Analysis of BSE 2024, EACVI/ASE 2016 and proposed algorithm in Predicting LV pre-A > 12 mmHg, LV pre-A >15 mmHg and LVEDP>15 mmHg for reduced ejection fraction.

|  |  |  |  |  |  |  |  |
| --- | --- | --- | --- | --- | --- | --- | --- |
| Pressure wave cut-off | Method | AUC | Sensitivity | Specificity | PPV | NPV | p-value |
| LV pre-A > 12 mmHg | BSE_2024 | 0.5714 | 0.1429 | 1.0000 | 1.0000 | 0.3793 | 1.2506 |
|  | EACVI_2016 | 0.6429 | 0.2857 | 1.0000 | 1.0000 | 0.4231 |  |
|  | Proposed | 0.8139 | 0.8095 | 0.8182 | 0.8947 | 0.6923 | Vs BSE<br>0.1249<br>vs EACVI<br>0.3466 |
| LV pre-A>15 mmHg | BSE_2024 | 0.5833 | 0.1667 | 1.0000 | 1.0000 | 0.4828 | 1.1038 |
|  | EACVI_2016 | 0.6667 | 0.3333 | 1.0000 | 1.0000 | 0.5385 |  |
|  | Proposed | 0.7738 | 0.8333 | 0.7143 | 0.7895 | 0.7692 | Vs BSE<br>0.2932<br>vs EACVI<br>0.7947 |
| LVEDP>15 mmHg | BSE_2024 | 0.5435 | 0.0870 | 1.0000 | 1.0000 | 0.1923 | 1.4765 |
|  | EACVI_2016 | 0.6087 | 0.2174 | 1.0000 | 1.0000 | 0.2174 |  |
|  | Proposed | 0.8913 | 0.7826 | 1.0000 | 1.0000 | 0.5000 | Vs BSE<br>0.0514<br>vs EACVI<br>0.1165 |

BSE = British Society of Echocardiography, EACVI = European Association of Cardiovascular Imaging, ASE: American Society of Cardiology. LVEDP: left ventricular end diastolic pressure. AUC: Area under the curve, NPV: negative predictive value, PPV: positive predictive value.

Additionally, agreement metrics in Table 11 supplementary (108 normal-normal, 19 elevated-elevated, 6 discordant) suggest that while BSE and EACVI are generally concordant, they still underestimate true hemodynamic burden revealed by the proposed enhanced method. This stronger NPV is critical in ruling out elevated pressures in ambiguous cases.

Also, when comparing ROC curves across Figures 1–9, visual analysis supports the tabulated data: the proposed algorithm consistently exhibits larger AUCs and more leftward-shifted curves than both BSE 2024 and EACVI/ASE 2016. Particularly, Figure 3 (ROC for LV pre-A ≥12 mmHg) and Figure 9 (ROC for LVEDP >15 mmHg) clearly illustrate the superior discriminative capacity of the proposed tool.

**Figure 1:**
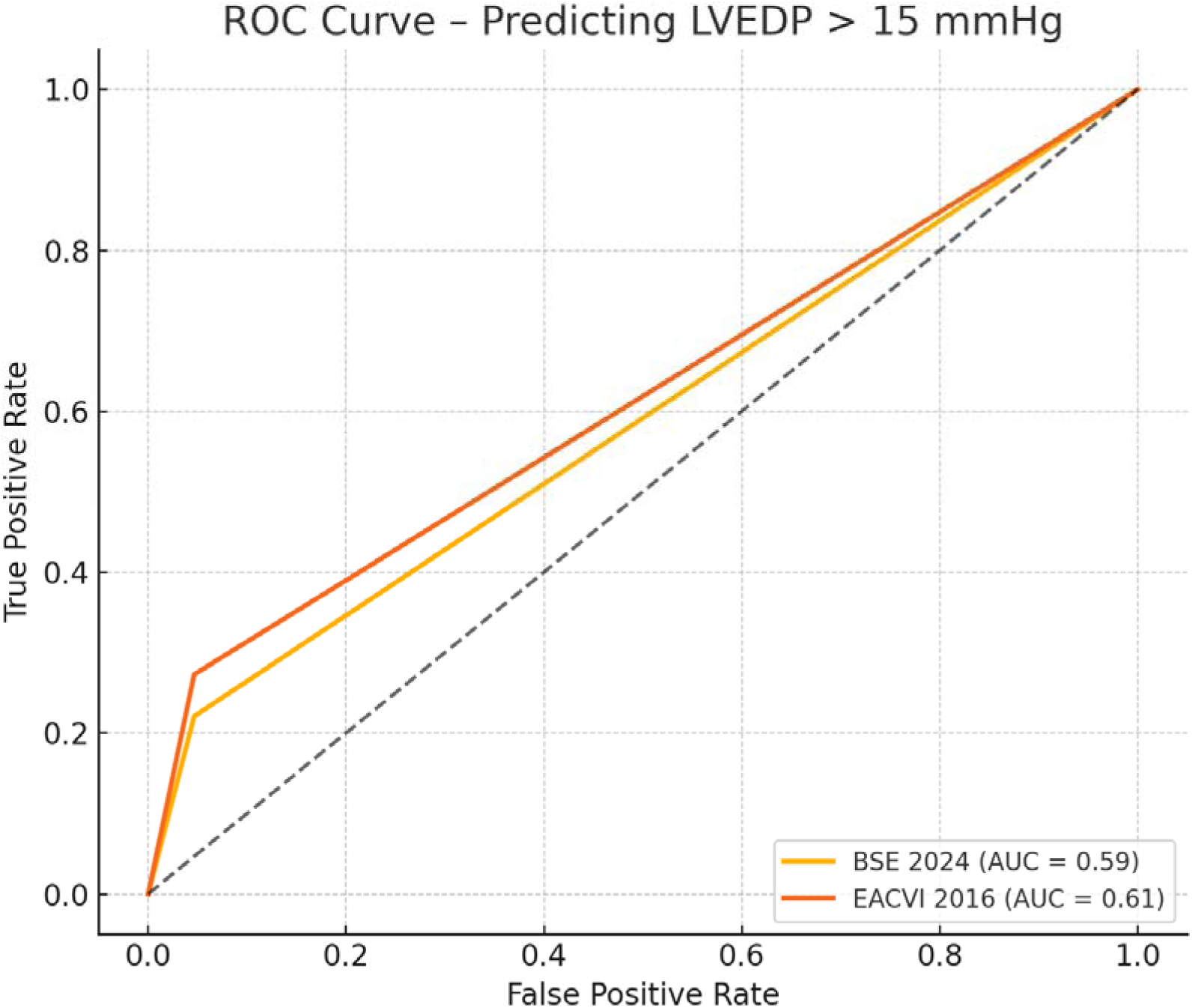
ROC Curve comparing BSE 2024 and EACVI 2016 pressure classifications against invasive LVEDP > 15 mmHg.

**Figure 2:**
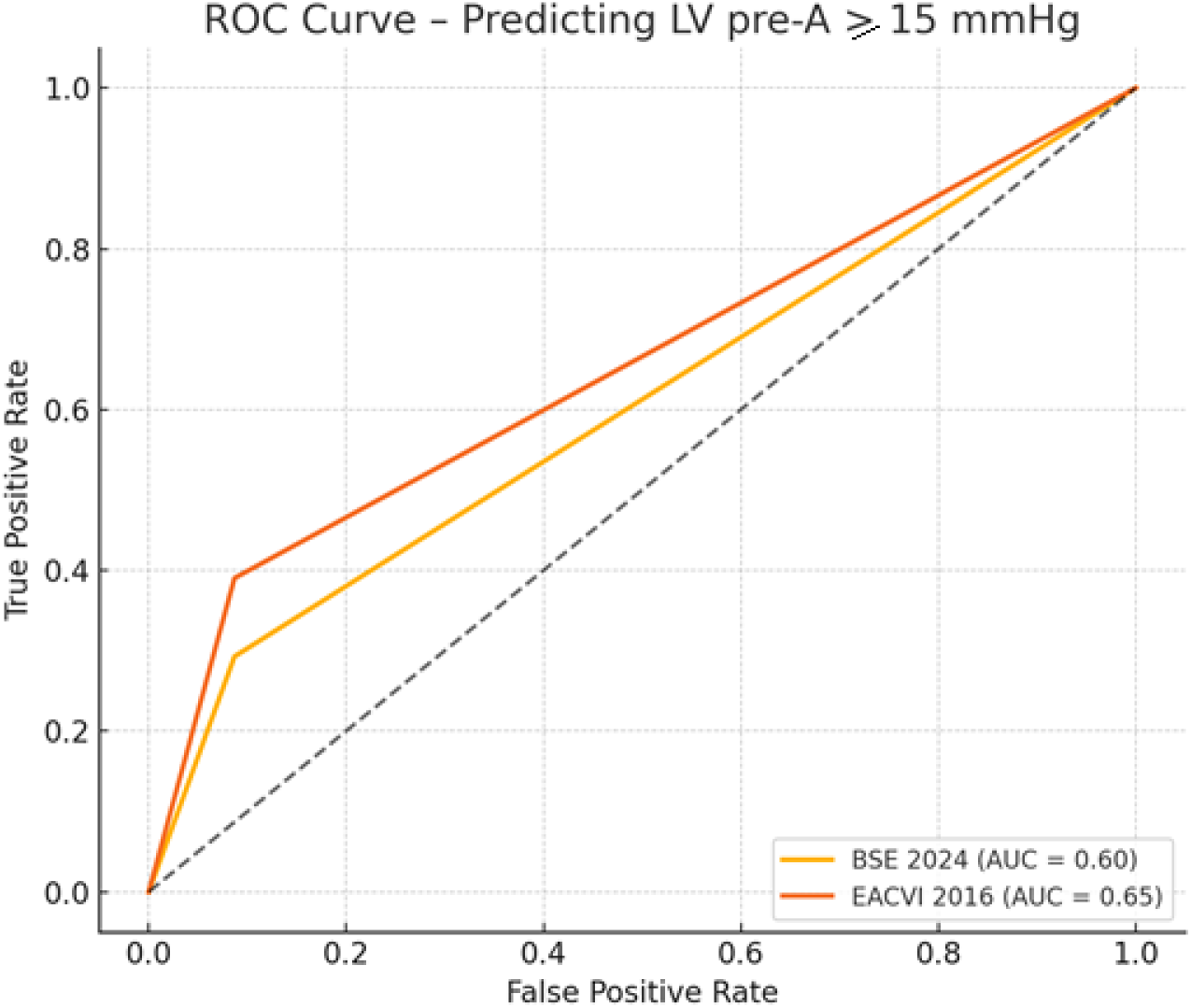
ROC Curve comparing BSE 2024 and EACVI 2016 pressure classifications against invasive LV pre-A ≥ 15 mmHg.

**Figure 3:**
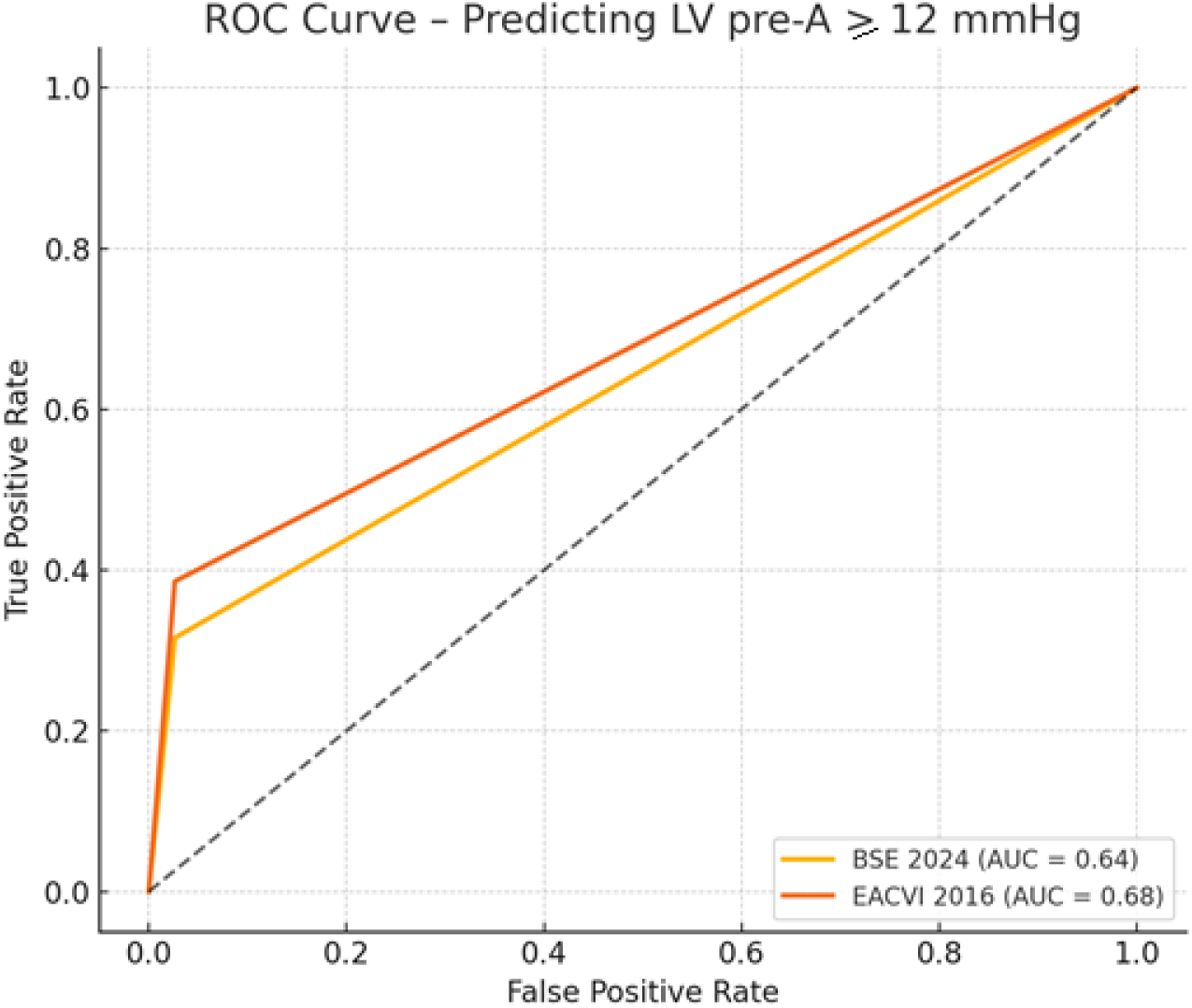
ROC Curve comparing BSE 2024 and EACVI 2016 pressure classifications against invasive LV pre-A ≥ 12 mmHg.

**Figure 4:**
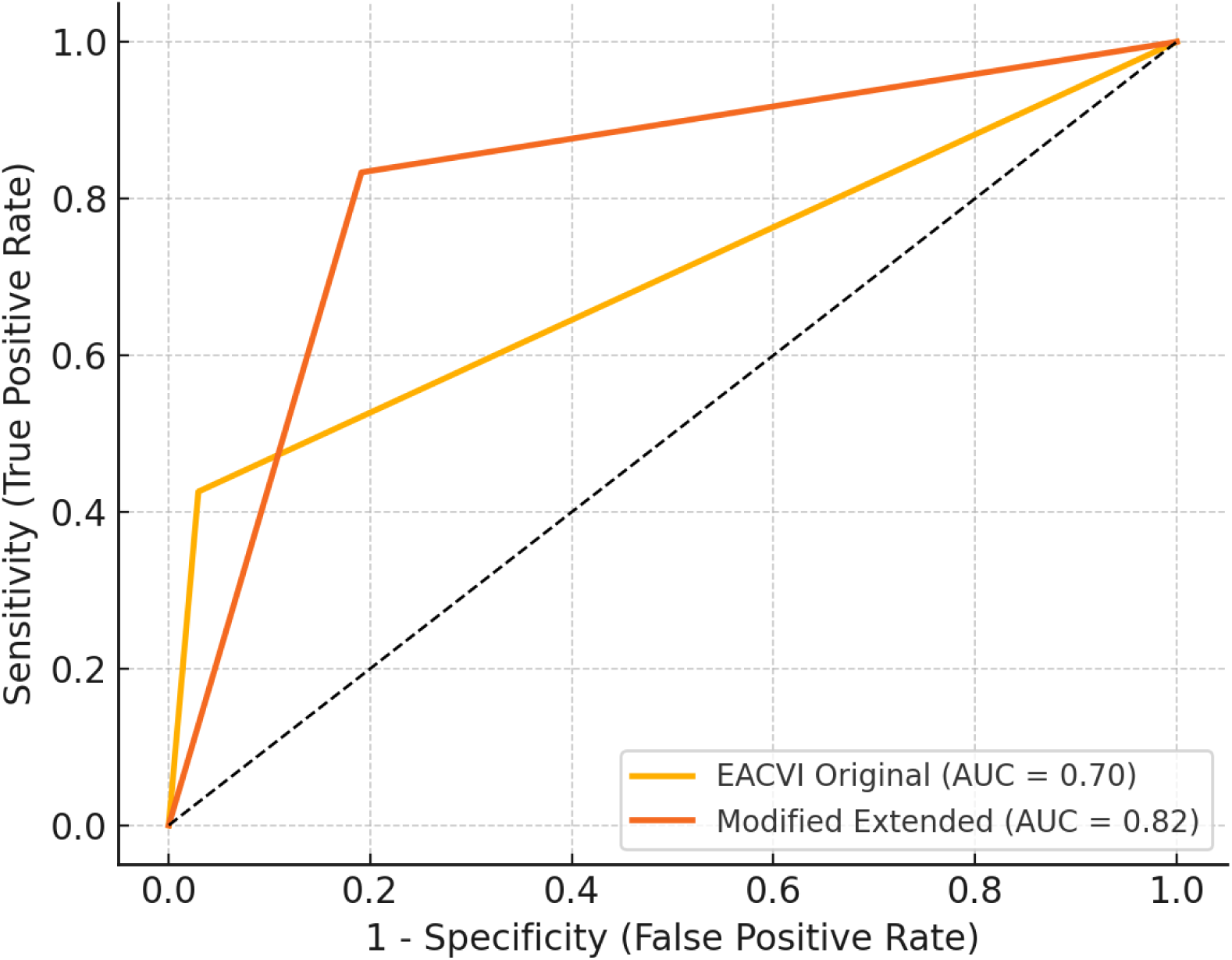
ROC Curve comparing EACVI Original and Modified Extended (LV pre-A ≥ 12 mmHg)

**Figure 5:**
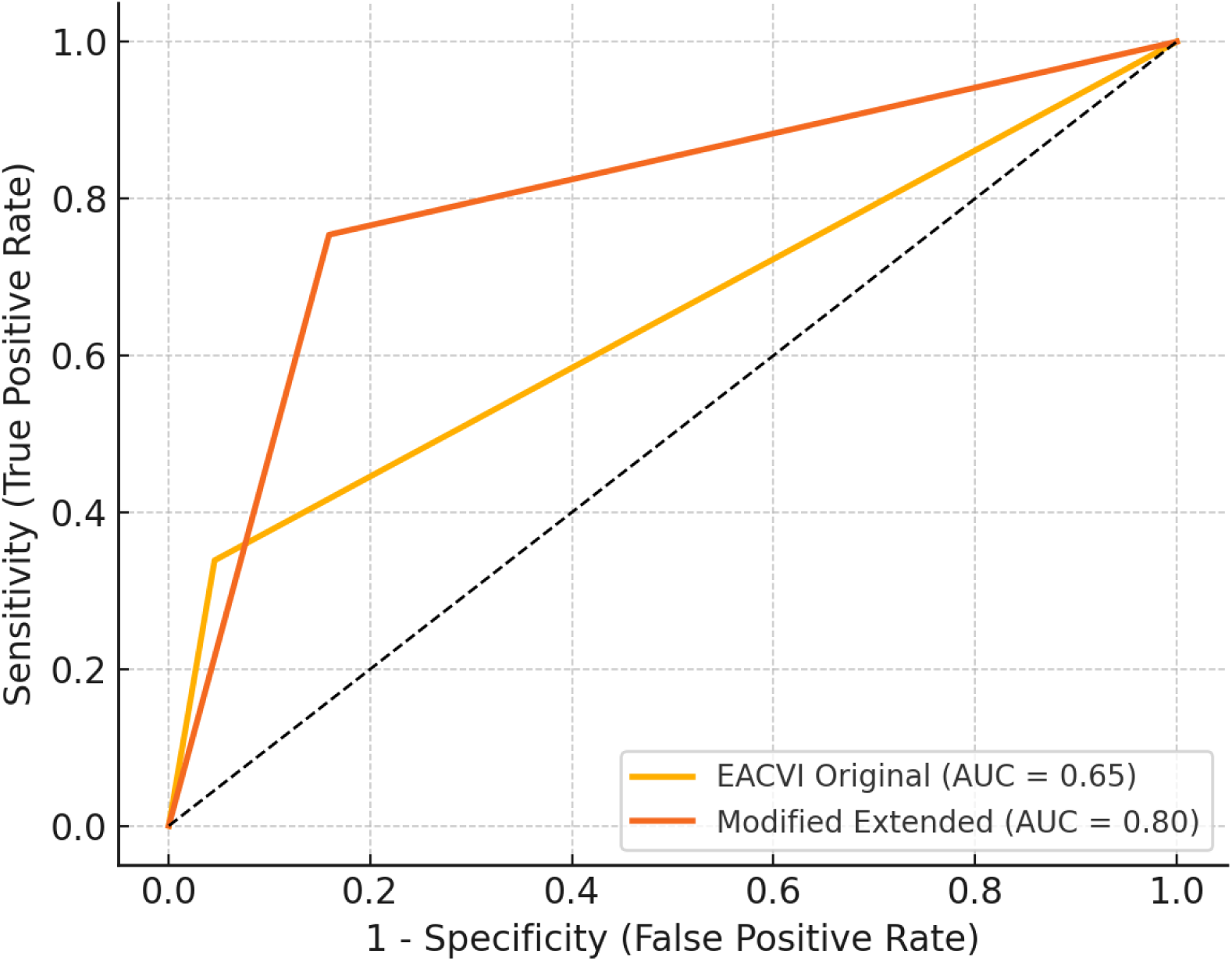
ROC Curve comparing EACVI Original and Modified Extended (LVEDP > 15 mmHg)

**Figure 6:**
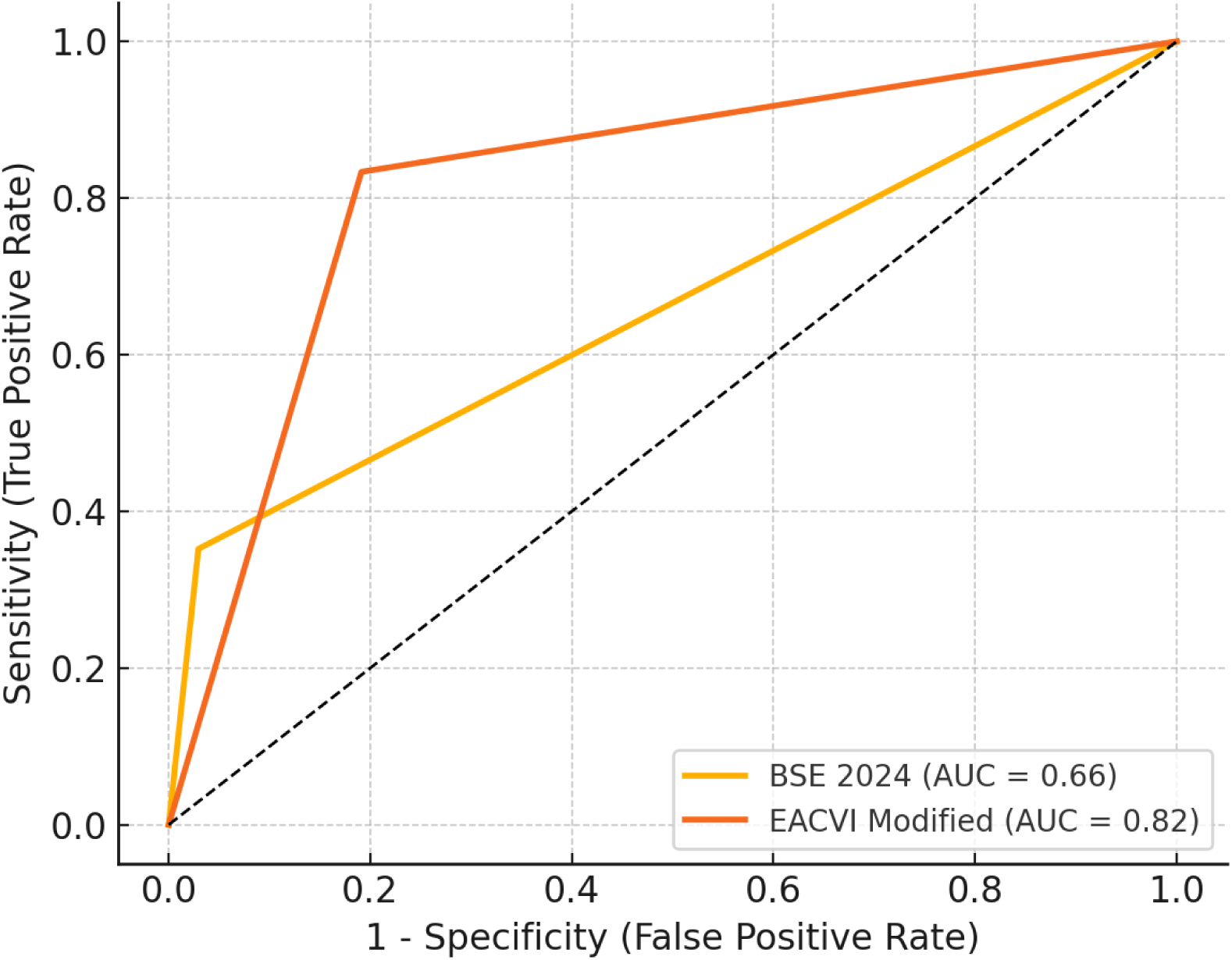
ROC Curve comparing BSE 2024 and EACVI Modified (Extended) for detecting elevated LV pre-A ≥12 mmHg.

**Figure 7.**
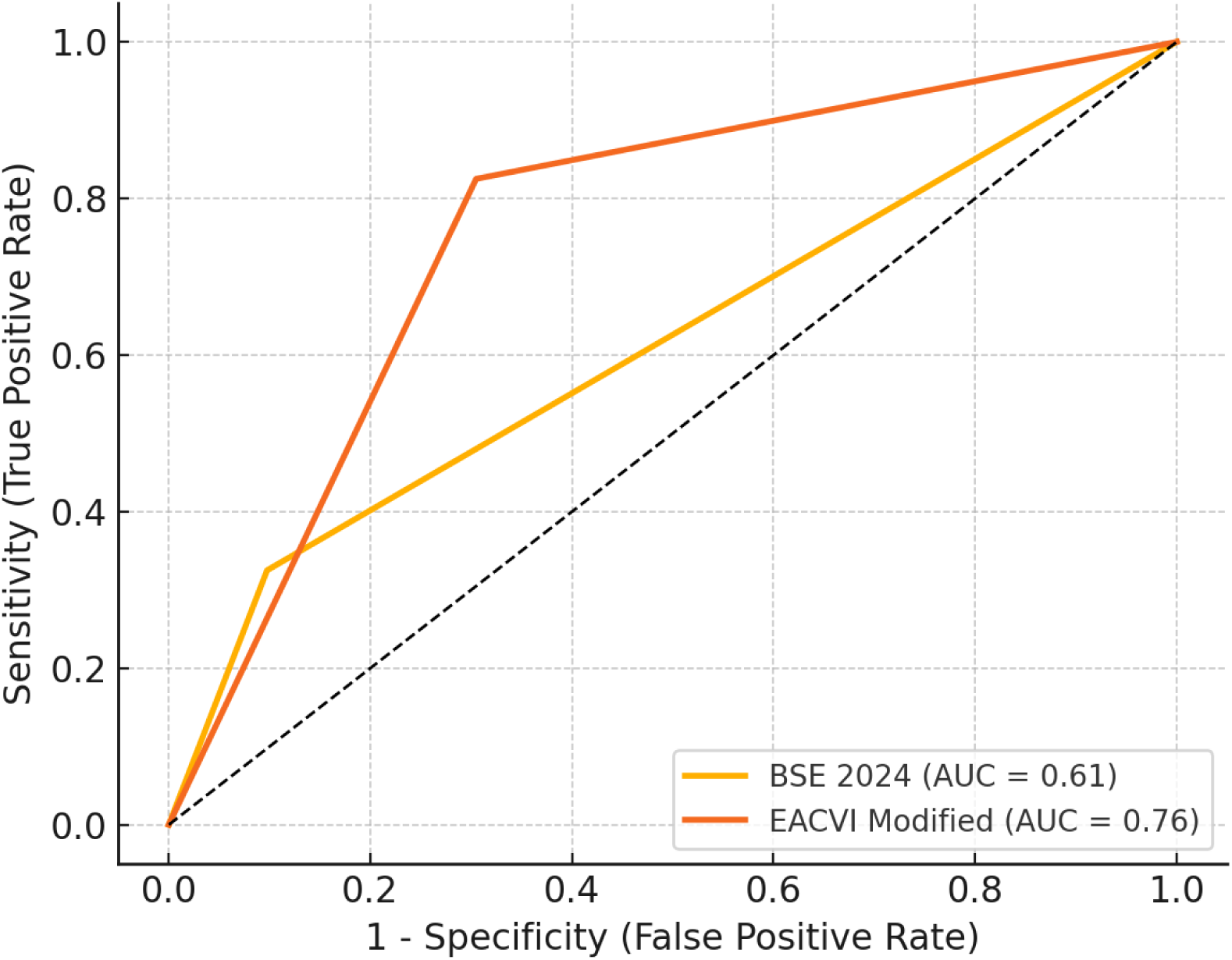
ROC Curve comparing BSE 2024 and EACVI Modified (Extended) for detecting elevated LV pre-A ≥15 mmHg.

**Figure 8:**
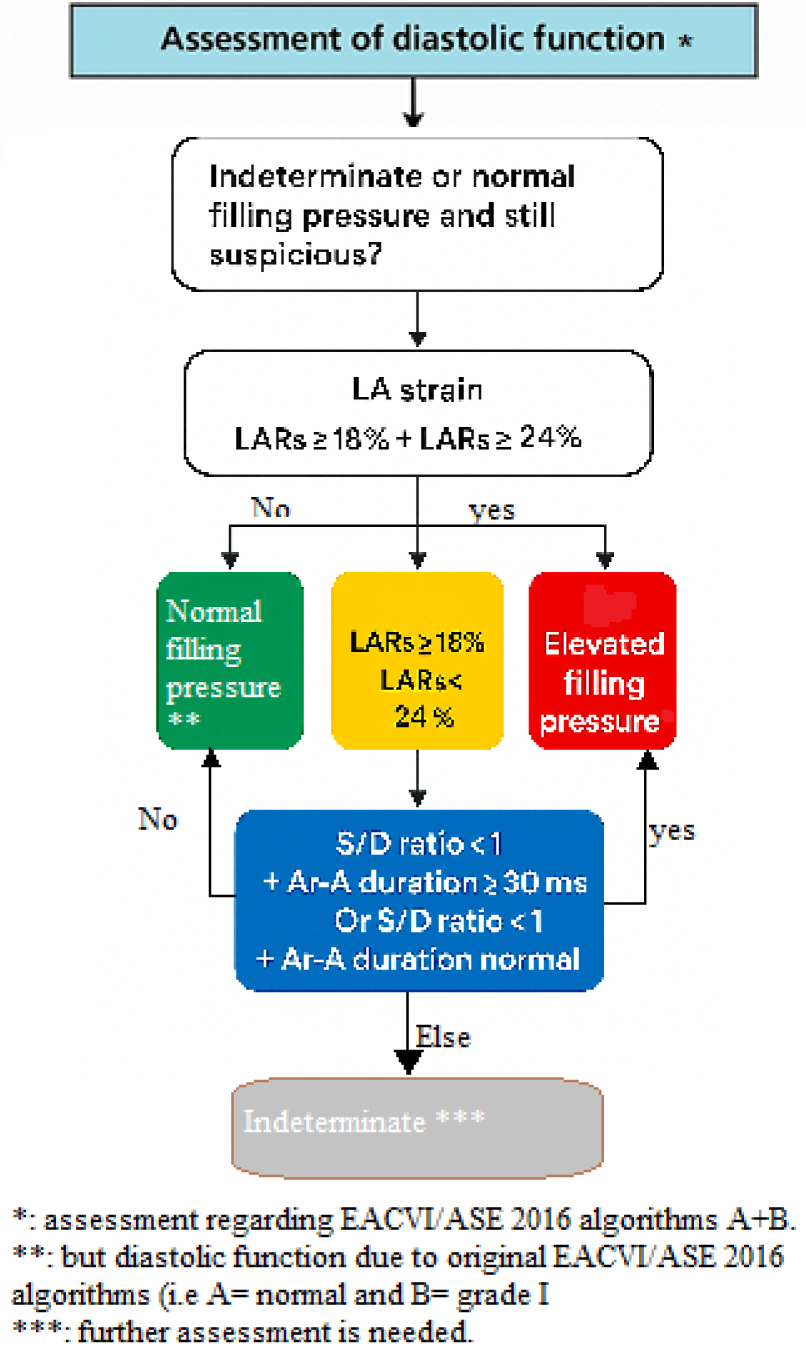
proposed algorithm for further assessment of LV filling pressure.

The utility of EACVI/ASE 2016 grading stratification was also partially validated, as mean LVEDP and pre-A increased across Grade I to Grade III (Table 9 supplementary). However, the lack of statistically significant pressure differences between adjacent grades (e.g., Grade II vs III: p=1.0 for LVEDP) calls into question the clinical utility of such granular classification and may explain why binary classification (normal vs impaired) performs better in ROC analysis.

Taken together, these supplemental findings affirm that while BSE and EACVI offer useful frameworks, a physiology-based, multi-parametric model—such as the one proposed—delivers more consistent, statistically robust, and clinically adaptable results across a range of patient phenotypes and invasive reference thresholds.

Additionally, strong agreement between BSE and EACVI in filling pressure classification (Cohen’s kappa = 0.837) highlights their general consistency, though divergence from the proposed algorithm underscores potential diagnostic underestimation in both current standards.

Recent advancements in diastolic function assessment have centered around LA strain parameters and their clinical integration. The global trend, especially between 2022–2025, has shifted from static metrics like LAVI and E/e′ toward dynamic functional markers. Notably, the Late Atrial Strain Evaluation (LATE) model [7] and the HFA-PEFF algorithm update in 2023 [8] recommend including LA reservoir strain (LARs) as a core criterion.

Comparative studies by Mavinkurve-Groothuis (2022) [7], Morris (2020) [9], and Inoue (2023) [10] have validated LARs against PCWP and LVEDP, with reported AUCs >0.80 in various HFpEF cohorts [11]. These study findings are consistent with this direction, showing the proposed algorithm achieving AUCs of 0.82 (LV pre-A ≥12 mmHg) and 0.80 (LVEDP >15 mmHg), outperforming both BSE 2024 [3] and EACVI/ASE 2016 [1] algorithms.

In this study, LV pre-A was found to be more stable and discriminative than LVEDP. This is in line with the notion that LVEDP is preload-sensitive and phasic, making LV pre-A a more reliable surrogate in ambulatory or borderline patients. These nuances have been emphasized in invasive validation papers by Andersen (2021) [12] and Verbrugge (2023) [13].

This study fills a critical gap by providing the first prospective comparison between EACVI 2016 and BSE 2024 [3] using invasive hemodynamics in a Middle Eastern cohort. These results may guide future regional adaptations or validations of diastolic function algorithms in similar populations.

### Clinical Implications

Incorporating LA strain and adjunctive parameters may significantly improve the diagnostic yield for diastolic dysfunction [14]. As echocardiography remains the cornerstone of HF diagnosis, enhancing its precision with newer tools is crucial for personalized patient care.

Although invasive pressure metrics such as LVEDP and LV pre-A are not prognostic endpoints per se, they remain the reference standard for validating noninvasive diastolic assessment tools. Their use in this study enabled a direct comparison between guideline-based algorithms and the proposed model against true hemodynamic burden.

By demonstrating superior diagnostic performance across multiple thresholds and invasive indices, the proposed algorithm shows strong potential to refine diastolic function classification.

This diagnostic refinement is not merely academic. More accurate classification of LV filling pressure may influence clinical decision-making regarding diuretic therapy, follow-up strategy, and need for invasive testing—particularly in patients with borderline findings or preserved EF.

Therefore, while not predictive of long-term outcomes directly, the accurate estimation of invasive pressures can support a higher level of individualized patient care, bridging the gap between physiology and practical cardiology.

Future multicenter studies across diverse populations are warranted to validate these findings and promote broader integration of advanced echocardiographic markers into guideline-based practice.

## 5. Conclusion

This prospective study provides the first comprehensive validation of the BSE 2024 [3] and EACVI/ASE 2016 [1] diastolic function algorithms against invasive hemodynamic standards in a Middle Eastern population. While both guidelines demonstrated moderate diagnostic performance, this study finds highlight the limitations of current criteria— particularly in sensitivity and in classifying ambiguous cases.

The novel algorithm proposed herein, integrating left atrial reservoir strain (LARs), pulmonary venous S/D ratio, and Ar–A duration, showed superior accuracy, higher predictive value, and consistency across ejection fraction subgroups. It also maintained discriminative power across both LVEDP and LV pre-A thresholds, emphasizing its robustness against variable physiological states.

The results affirm the global trend toward a physiology-based, multiparametric approach in diastolic function assessment. Incorporating advanced functional markers such as LA strain into routine evaluation may significantly improve diagnostic precision, risk stratification, and patient management.

Future multicenter studies across diverse populations are warranted to confirm these findings and to encourage the evolution of clinical guidelines toward dynamic, evidence-based frameworks.

## 6. Limitations and Future Directions

This study has several important limitations. First, it was conducted at a multicenter with a relatively homogenous Middle Eastern population, which may limit generalizability to other ethnic and geographic groups. Second, patients with atrial fibrillation, significant valvular disease, or suboptimal image quality were excluded, potentially introducing selection bias.

Additionally, LA strain analysis was performed using vendor-specific software, which may limit reproducibility across different echocardiographic platforms.

Despite these limitations, the study provides compelling evidence for incorporating multiparametric indices, especially LA strain and pulmonary venous flow, into diastolic function assessment frameworks.

Future research should focus on multi-center, multi-ethnic validation of the proposed algorithm, incorporating patients with broader clinical characteristics including arrhythmias, valvular pathology, and HF with preserved ejection fraction.

Randomized clinical trials assessing clinical outcomes based on algorithm-driven classification may also clarify the prognostic and therapeutic implications of refined diastolic assessment.

## Conflict of Interest

The authors declare that they have no conflict of interest relevant to the content of this manuscript.

## Funding

This research received no specific grant from any funding agency in the public, commercial, or not-for-profit sectors.

## Data Availability

I hereby confirm the availability of all data on asking

## Notes

### Competing Interest Statement

The authors have declared no competing interest.

### Funding Statement

No funding received

### Author Declarations

The approval from the Damascus University ethics committe was granted for a master degree thesis

