## Supplementary material for "Comparative Validation of BSE and EACVI/ASE Guidelines for Estimating Filling Pressure: Proposal of an Algorithm": Supllementry tables 9 to 13

*.*

| Table 9: Diastolic function classification Cross-Tabulation | | |
| --- | --- | --- |
| BSE \ EACVI | EACVI Normal | EACVI Impaired |
| BSE Normal | 12 | 0 |
| BSE Impaired | 67 | 54 |

*Analysis includes only patients with definitive classification (Normal or Impaired) by both guidelines.
BSE = British Society of Echocardiography 2024; EACVI = European Association of Cardiovascular Imaging, ASE : American Society of Cardiology.*

| Table 10: EACVI Diastolic Grades vs Invasive LV pre-A and LVEDP (Continuous) | | | | |
| --- | --- | --- | --- | --- |
| EACVI Diastolic Grade | Mean LV pre-A ± SD (mmHg) | P value comparison same line grade vs next line grade | Mean LVEDP± SD (mmHg) | P value comparison same line grade vs next line grade |
| Indeterminate | 17.02 ± 1.00 | p=0.0444 | 23.62 ± 2.29 | p=0.2113 |
| Normal | 10.16 ± 4.22 | p=0.0009 | 16.24 ± 5.56 | p=0.0006 |
| Grade I | 14.27 ± 4.98 | p=0.6524 | 22.58 ± 8.42 | p=0.9268 |
| Grade II | 16.24 ± 5.05 | p=0.0744 | 24.26 ± 8.39 | p=1.0 |
| Grade III | 21.56 ± 8.43 | NA | 24.00 ± 7.07 | NA |

*EACVI = European Association of Cardiovascular Imaging, ASE : American Society of Cardiology. LVEDP: left ventricular end diastolic pressure.*

| Table 11: Comparison of BSE 2024 and EACVI/ASE 2016 Filling Pressure Classifications | | |
| --- | --- | --- |
| Category | BSE 2024 (n, %) | EACVI 2016 (n, %) |
| Elevated Pressure | 20 (15.0%) | 24 (18.0%) |
| Normal Pressure | 113 (85.0%) | 109 (82.0%) |

*BSE = British Society of Echocardiography, EACVI = European Association of Cardiovascular Imaging, ASE: American Society of Cardiology*

| Table 12: Agreement Between BSE 2024 and EACVI/ASE 2016 in Filling Pressure Classification Cross-Tabulation:*.* | | | | |
| --- | --- | --- | --- | --- |
| BSE 2024 / EACVI_ASE 2016 | Normal | Elevated | Cohen's Kappa | McNemar's Test P-value |
| Normal | 108 | 5 | 0.837 | 0.221 |
| Elevated | 1 | 19 |  |  |

*BSE = British Society of Echocardiography, EACVI = European Association of Cardiovascular Imaging, ASE: American Society of Cardiology..*

| Table 13: ROC Analysis of BSE 2024, EACVI/ASE 2016 and proposed algorithm in Predicting LV pre-A > 12 mmHg, LV pre-A>15 mmHg and LVEDP>15 mmHg for preserved ejection fraction | | | | | | | |
| --- | --- | --- | --- | --- | --- | --- | --- |
| Pressure wave cut-off | Method | AUC | Sensitivity | Specificity | PPV | NPV | p-value |
| LV pre-A > 12 mmHg | BSE_2024 | 0.6862 | 0.4074 | 0.9649 | 0.8462 | 0.7746 | 2.0000 |
|  | EACVI_2016 | 0.6862 | 0.4074 | 0.9649 | 0.8462 | 0.7746 |  |
|  | Proposed | 0.7729 | 0.7037 | 0.8421 | 0.6786 | 0.8571 | Vs BSE or EACVI 0.6423 |
| LV pre-A>15 mmHg | BSE_2024 | 0.6490 | 0.3889 | 0.9091 | 0.5385 | 0.8451 | 2.0000 |
|  | EACVI_2016 | 0.6490 | 0.3889 | 0.9091 | 0.5385 | 0.8451 |  |
|  | Proposed | 0.6768 | 0.6111 | 0.7424 | 0.3929 | 0.8750 | Vs BSE or EACVI 1.5951 |
| LVEDP>15 mmHg | BSE_2024 | 0.6271 | 0.3056 | 0.9487 | 0.8462 | 0.5968 | 2.0000 |
|  | EACVI_2016 | 0.6271 | 0.3056 | 0.9487 | 0.8462 | 0.5968 |  |
|  | Proposed | 0.7415 | 0.6111 | 0.8718 | 0.8148 | 0.7083 | Vs BSE or EACVI  0.3746 |

*BSE = British Society of Echocardiography, EACVI = European Association of Cardiovascular Imaging, ASE: American Society of Cardiology. LVEDP: left ventricular end diastolic pressure. AUC: Area under the curve, NPV: negative predictive value, PPV: positive predictive value..*
